# Herpes Simplex Virus Serostatus and 1-Year Mortality Among Allogeneic Hematopoietic Cell Transplant Recipients

**DOI:** 10.64898/2026.07.30.26359369

**Authors:** Molly D. Fischer, Christine Johnston, Michael J. Boeckh, Emily S. Ford, Ted Gooley, Amanda I. Phipps, Rachel L. Winer, Melinda A. Biernacki, Denise J. McCulloch, Brenda M. Sandmaier, Alex L. Greninger, Anna Wald, Steven A. Pergam

## Abstract

Viral infections remain a cause of substantial morbidity and mortality in allogeneic hematopoietic cell transplant (aHCT) recipients. While antiviral prophylaxis has dramatically reduced the risk of herpes simplex virus (HSV) disease, the relationship between HSV serostatus and major post-transplant complications in the context of HSV prophylaxis is unknown. We evaluated the association between HSV serostatus and survival among adults who received a first aHCT at the Fred Hutchinson Cancer Center between 2002 and 2022. Patients were screened for HSV-1 and HSV-2 by Western blot (WB) prior to transplant. We fit Cox proportional hazards models for mortality up to one year post-transplant, comparing HSV seropositive to seronegative patients. Models were adjusted for age, sex, cytomegalovirus (CMV) serostatus, conditioning regimen, disease risk, graft type and HLA matching, year of transplant and acute graft-versus host disease. A total of 4,016 aHCT recipients were included in this analysis. The cumulative all-cause 1-year mortality was 29.8%. For HSV-1, the adjusted hazard ratio (aHR) for all-cause mortality comparing seropositive to seronegative individuals was 1.20 (95% CI: 1.06-1.36). The aHRs for relapse, non-relapse mortality (NRM) and relapse-related mortality (RRM) were 1.46 (1.24-1.71), 1.04 (0.89-1.21), and 1.75 (1.40-2.19), respectively. For HSV-2, the aHRs for all-cause mortality, relapse, NRM, and RRM were 1.03 (0.93-1.14), 1.16 (1.03-1.31), 0.99 (0.87-1.13), and 1.13 (0.97-1.33), respectively. Despite universal antiviral prophylaxis, HSV-1 seropositivity was associated with higher mortality in the year after transplant, driven by RRM. Further studies are needed to confirm the association and understand the potential mechanisms underlying this relationship.

**Key Points:**

- Pre-transplant HSV-1 seropositivity was associated with higher 1-year mortality among adult aHCT recipients over a 20-year period
- HSV seropositivity was associated with relapse and relapse-related mortality; seropositivity was high in the study population

## Introduction

Survival among persons undergoing allogeneic hematopoietic cell transplant (aHCT) has improved over the past 30 years; however, infection and inflammatory complications remain the most common causes of non-relapse mortality.^1,2^ Due to profound immunosuppression, aHCT recipients are at risk for complications resulting from reactivation of several human herpesviruses, including cytomegalovirus (CMV),^3–6^ human herpesvirus 6,^7^ Epstein-Barr virus (EBV),^8^ and herpes simplex virus types 1 and 2 (HSV-1/2).^9–11^ Prior to the implementation of HSV prophylaxis using acyclovir (ACV) or valacyclovir (VACV), most transplant recipients developed HSV mucocutaneous diseases,^9^ and some developed severe HSV-related complications including hepatitis,^10^ pneumonia,^11^ and ACV-resistant HSV. The use of HSV prophylaxis in the post-transplant period dramatically reduces the risk of both mild and severe clinical disease.^12^ However, prophylaxis does not completely eliminate subclinical (or asymptomatic) HSV reactivation. HSV shedding, detected by PCR of mucosal swabs, occurs in immunocompetent patients even during high-dose antiviral prophylaxis,^13^ and asymptomatic HSV-1 shedding both on and off ACV has been documented among aHCT patients and other immunosuppressed patient populations.^14–18^

Recipient CMV seropositivity has been associated with increased mortality,^4–6^ despite overall reductions in clinical CMV disease as a result of pre-emptive treatment with agents such as ganciclovir. More recently, letermovir prophylaxis has been shown to decrease the risk of CMV reactivation post-transplant,^19^ and may also lower the risk of mortality beyond day 200 post-transplant.^20^ Whether there is a similar adverse mortality impact related to HSV serostatus in the setting of universal prophylaxis is unknown. Because of the high population-level seroprevalence of HSV, widespread use of anti-HSV prophylaxis, and false positive or false negative results on some FDA-authorized binding HSV serologic assays,^21,22^ not all transplant centers routinely assess recipient HSV serostatus prior to transplant. Furthermore, among centers that do screen prior to aHCT, the Fred Hutchinson Cancer Center in Seattle, WA is one of the only centers that uses the University of Washington Western blot (WB), which is considered the gold standard HSV serologic assay.^23^

The objective of this study was to evaluate the association between pre-transplant HSV-1 and HSV-2 serostatus and mortality in the 1-year post-transplant period to understand whether, similar to CMV, HSV infection may influence post-transplant outcomes beyond HSV disease. We hypothesized that despite prophylaxis, HSV seropositive aHCT recipients would be at higher risk for all-cause mortality than seronegative recipients, driven by a difference in non-relapse mortality (NRM).

## Methods

### Study Population

Using a center-specific transplant database,^1^ we reviewed data from all adults (age ≥ 18) who received their first aHCT at the Fred Hutchinson Cancer Center (FHCC) between January 1, 2002, and December 31, 2022. Genetically identical (iso) transplant recipients and those who received a prior aHCT outside of FHCC were excluded. The date of last follow-up before the dataset was locked was April 26, 2024. This study was approved by the Institutional Review Board at FHCC under Protocol 1829 (Substudy 060).

### Transplantation Methods, Conditioning, GVHD Prophylaxis

Detailed descriptions of transplantation methods over the study period have been described in prior publications.^1,24^ Prior to transplantation, all patients received a conditioning regimen that was classified as either low, intermediate, or high intensity using the transplant conditioning intensity (TCI) score^25^ combined with good clinical judgment.^26^ GVHD prophylaxis generally consisted of calcineurin inhibitors (CNI) plus methotrexate (MTX) or mycophenolate mofetil (MMF) with or without sirolimus (SIRO); alternatively, post-transplant cyclophosphamide (PTCy) was used alone or combined with one or more of the above-listed agents.^24^ Regimens were categorized into one of the following groups based on the “backbone” of the regimen: CNI only, CNI plus MMF, CNI plus MMF plus SIRO, CNI plus MTX, CNI plus SIRO, SIRO plus MMF, PTCy, or a clinical trial regimen.

### Herpesvirus Serology and Prophylaxis

Type-specific HSV serostatus is routinely assessed for all patients prior to transplant at FHCC using the University of Washington WB;^23^ donor HSV serostatus is not routinely assessed. Serology results were classified as seropositive, seronegative, indeterminate or uninterpretable for HSV-1 and for HSV-2. Patients with missing, indeterminate or uninterpretable HSV-1/2 serostatus were excluded from the primary analyses. Both donor and recipient CMV serostatus are routinely assessed pre-transplant by FDA-approved CMV assays. VZV/HSV prophylaxis consisted of ACV 250 mg/m^2^ administered intravenously during periods when patients could not tolerate oral medications, and transitioned to either 800mg ACV or 500mg VACV orally twice daily for at least one year post-transplant.^12,27,28^ CMV-seropositive cord blood recipients received high-dose (2g three times daily) VACV prophylaxis for CMV prevention from June 2008 until October 2018, after which routine letermovir prophylaxis was implemented at FHCC.^29^

### Statistical Analyses

The primary outcome of interest was all-cause mortality within one year of transplant. We fit Kaplan-Meier curves to model the unadjusted survival function over time comparing seropositive to seronegative patients for HSV-1, HSV-2, or CMV as a comparator. We then fit Cox proportional hazards models to compare time to all-cause mortality between seropositive and seronegative aHCT recipients, adjusting for the factors described below and truncating follow-up at one-year post-transplant. We also evaluated cause-specific mortality by constructing cumulative incidence functions for relapse, death preceded by relapse (RRM), and death not preceded by relapse (NRM). Cause-specific Cox proportional hazards were fit for all outcomes, with observations censored at the time of NRM for the outcomes of relapse and RRM, and at the time of relapse for the outcome of NRM. For the cause-specific analyses, patients with aplastic anemia, solid tumors, and nonmalignant conditions were excluded due to differences in the risk and biological process of relapse between these and malignant conditions. Patients who received a subsequent aHCT within 1 year of first aHCT were censored at the time of subsequent transplant for the cause-specific survival analyses. We further evaluated mortality within cohorts defined by transplant year (early: 2002-2012 and late: 2013-2022) to understand how incremental changes in transplant protocols over time may have impacted this association. Patient characteristics were compared between cohorts by chi square tests for categorial variables and Wilcoxon rank test for continuous variables. Analyses were conducted in RStudio version 2024.09.1+394.^30^

### Covariates

Models were adjusted for covariates selected *a priori* as factors associated with risk of post-transplant mortality, including recipient age, sex, underlying disease risk (including disease, stage, and cytogenetics where applicable and available),^24,31^ recipient CMV serostatus, conditioning intensity, graft type and HLA matching, and year of transplant to account for potential changes over time in transplant practices (see **Supplemental Table 1** for further description of covariates and **Supplemental Figure 1** for a directed acyclic graph). Acute graft-versus-host disease (aGVHD) grade II or higher was also included as a time-varying covariate in the model. The FHCC grading system for aGVHD was updated for patients who received transplants in 2022 and later, in which grade II is divided into IIa and IIb. A grade of IIa or higher was considered equivalent to grade II or higher for this analysis.

### Sensitivity Analyses

To assess the robustness of our findings and explore the roles of potential confounders, we conducted several sensitivity analyses. First, we assessed the potential interactions between CMV and HSV serostatus, and conditioning regimen and HSV serostatus by fitting models with and without interaction terms and determined whether model fit was improved by ANOVA. Second, we evaluated post-transplant CMV reactivation (defined as any positive CMV test result (ex: PCR, antigen test) within the first year) as a time-varying covariate into our models. Third, due to the possibility that aGVHD may lie on the causal pathway between HSV-1 serostatus and mortality, we constructed additional models excluding aGVHD. Fourth, we investigated the potential impacts of including patients with indeterminate HSV serostatus on our estimates. Since clinically persons with indeterminate HSV serostatus results are treated as seronegative, we included those with indeterminate HSV values and classified them as seronegative.

We also conducted an exploratory analysis to evaluate whether there was an association between pre-transplant blood transfusions and HSV and CMV serostatus. For patients who had a transplant between 2020-2022, ICD-10 billing codes were used to identify any blood transfusions received in the 90 days prior to HSV WB testing. Transfusions received on the day of WB testing were excluded, as the temporality in relation to testing was unknown. We fit univariate and multivariable logistic regression models to examine whether the number of blood transfusions was associated with odds of HSV and CMV seropositivity.

## Results

### Patient Demographics

A total of 4,381 patients received their first aHCT at FHCC between 2002-2022. After excluding 19 iso transplant recipients and 384 patients with missing, indeterminate or uninterpretable HSV serostatus, 4,016 aHCT recipients were included in this analysis. Demographic and transplant data for these patients are presented in **Table 1**. A detailed description of underling malignancy categories is included in **Supplemental Table 2**. The median age at transplant was 53 (range: 18-80 years) and 2,321 (57.8%) patients were male. Most patients (n = 3,276, 81.6%) were HSV-1 seropositive and 1,136 (28.3%) were HSV-2 seropositive; in total, 848 (21.1%) were seropositive for both HSV-1 and HSV-2. In the early (2002-2012) cohort, patients were younger (median age 51 vs. 56, p < 0.01) and a higher proportion had high-risk diseases (37.3% vs 21.2%, p < 0.001). Sixty-two patients with NHL, 108 patients with plasma cell neoplasms, 13 patients with Hodgkin’s disease, and 3 patients with CLL or MDS/MPN underwent tandem transplantation with myeloablative autograft followed within 3 months with reduced-intensity conditioning (or nonmyeloablative or both) allograft. HSV seropositivity was lower in the early cohort compared to the late cohort, with a greater difference observed for HSV-2 (24.1% vs. 33.8%, p < 0.01) than HSV-1 (80.6% vs. 82.8%, p = 0.08). CMV seropositivity was also slightly lower in the early (57.7%) vs. late (60.7%) cohort (p = 0.06). The age-stratified HSV and CMV serostatus distribution for the study population is shown in **Table 2**, alongside national HSV and CMV seroprevalence estimates.

**Table 1:** Patient Characteristics: Overall vs. Early (2002-2012) vs. Late (2013-2022) Cohorts.

|  | Overall<br>(N=4016) | Early<br>(N=2281) | Late<br>(N=1735) |
| --- | --- | --- | --- |
| **Age** |  |  |  |
| Median [Min, Max] | 53.5 [18.5, 80.9] | 51.3 [18.5, 76.1] | 56.8 [18.6, 80.9] |
| **Sex** |  |  |  |
| Male | 2320 (57.8%) | 1338 (58.7%) | 982 (56.6%) |
| Female | 1696 (42.2%) | 943 (41.3%) | 753 (43.4%) |
| **Underlying Malignancy** |  |  |  |
| ALL | 420 (10.5%) | 211 (9.3%) | 209 (12.0%) |
| AML | 1486 (37.0%) | 809 (35.5%) | 677 (39.0%) |
| Aplastic Anemia/PNH | 98 (2.4%) | 42 (1.8%) | 56 (3.2%) |
| CLL | 123 (3.1%) | 104 (4.6%) | 19 (1.1%) |
| CML | 196 (4.9%) | 147 (6.4%) | 49 (2.8%) |
| Hodgkin's Lymphoma | 91 (2.3%) | 71 (3.1%) | 20 (1.2%) |
| MDS/MPN | 985 (24.5%) | 469 (20.6%) | 516 (29.7%) |
| Non-Hodgkin's Lymphoma | 348 (8.7%) | 250 (11.0%) | 98 (5.6%) |
| Nonmalignant | 29 (0.7%) | 10 (0.4%) | 19 (1.1%) |
| Other Malignant | 30 (0.7%) | 21 (0.9%) | 9 (0.5%) |
| Plasma Cell Neoplasms | 210 (5.2%) | 147 (6.4%) | 63 (3.6%) |
| **Donor/HLA Matching** |  |  |  |
| HLA-Matched Sibling | 1213 (30.2%) | 818 (35.9%) | 395 (22.8%) |
| Mismatched Sibling or Non-Sibling<br>Relative | 253 (6.3%) | 133 (5.8%) | 120 (6.9%) |
| Unrelated | 2550 (63.5%) | 1330 (58.3%) | 1220 (70.3%) |
| **Tandem Auto-Allo Transplant** |  |  |  |
| No | 3830 (95.4%) | 2147 (94.1%) | 1683 (97.0%) |
| Yes | 186 (4.6%) | 134 (5.9%) | 52 (3.0%) |
| **Conditioning Intensity** |  |  |  |
| High | 2244 (55.9%) | 1373 (60.2%) | 871 (50.2%) |
| Intermediate | 655 (16.3%) | 168 (7.4%) | 487 (28.1%) |
| Low | 1117 (27.8%) | 740 (32.4%) | 377 (21.7%) |
| **Disease Risk** |  |  |  |
| Low | 330 (8.2%) | 143 (6.3%) | 187 (10.8%) |
| Intermediate | 2353 (58.6%) | 1245 (54.6%) | 1108 (63.9%) |
| High | 1229 (30.6%) | 850 (37.3%) | 379 (21.8%) |
| Very High | 48 (1.2%) | 0 (0%) | 48 (2.8%) |
| Unknown | 56 (1.4%) | 43 (1.9%) | 13 (0.7%) |
| **CMV Serostatus** |  |  |  |
| Seronegative | 1606 (40.0%) | 952 (41.7%) | 654 (37.7%) |
| Seropositive | 2369 (59.0%) | 1316 (57.7%) | 1053 (60.7%) |
| Missing | 41 (1.0%) | 13 (0.6%) | 28 (1.6%) |
| **HSV-1 Serostatus** |  |  |  |
| Seronegative | 740 (18.4%) | 442 (19.4%) | 298 (17.2%) |
| Seropositive | 3276 (81.6%) | 1839 (80.6%) | 1437 (82.8%) |
| **HSV-2 Serostatus** |  |  |  |
| Seronegative | 2880 (71.7%) | 1731 (75.9%) | 1149 (66.2%) |
| Seropositive | 1136 (28.3%) | 550 (24.1%) | 586 (33.8%) |

**Table 2:** Age-stratified CMV and HSV seroprevalence in the study population compared to NHANES estimates.

|  |  | FHCC |  |  | NHANES* |  |  |
| --- | --- | --- | --- | --- | --- | --- | --- |
|  |  | % Seropositive |  |  | % Seropositive (95% CI) |  |  |
| Age | N Total | CMV | HSV-1 | HSV-2 | CMV | HSV-1 | HSV-2 |
| 18-29 | 379 | 54.0 | 79.7 | 16.4 | 49.5 (46.1-52.8) | 41.3 (33.8-49.1) | 7.6 (4.7-11.5) |
| 30-39 | 524 | 58.3 | 82.1 | 21.4 | 56.7 (53.2-60.1) | 54.1 (48.6-59.5) | 13.3 (9.1-18.4) |
| 40-49 | 737 | 58.7 | 80.3 | 28.5 | 58.0 (54.8-61.1) | 59.7 (51.7-57.5) | 21.2 (16.3-26.8) |
| 50-59 | 1160 | 61.0 | 80.7 | 30.8 | - | - ** | - |
| 60-69 | 1,014 | 61.3 | 83.0 | 33.1 | - | - | - |
| 70-89 | 202 | 60.5 | 86.0 | 29.2 | - | - | - |

\* HSV data obtained from the 2015-2016 National Health and Nutrition Examination Survey conducted among individuals aged 14-49 in the United States.<sup>32</sup> CMV data obtained from the 1999-2004 NHANES cycle.<sup>34</sup> In this table, the population of aHCT patients at FHCC aged 18-29 is compared to the population aged 20-29 in NHANES.

### 1-Year Mortality and Other Post-Transplant Outcomes

In total, 1,195 patients (29.8%) died from any cause within one year of transplant. The 1-year cumulative mortality was significantly lower among patients who had a more recent transplant (2013-2022; 24.8%) compared to those who transplanted before 2012 (33.5%) (p < 0.01). The proportion of patients who relapsed within 1 year of transplant was similar between 2002-2012 (19.5%) and 2013-2022 (20.6%) (p = 0.4). The proportion of patients who developed aGVHD grade 2 or higher was lower in the later cohort (68.2% vs. 64.5%, p = 0.01).

### HSV and CMV Seropositivity and All-Cause Mortality

Overall, between 2002-2022, the adjusted hazard for all-cause mortality comparing HSV-1 seropositive to seronegative patients was 1.20 (95% CI: 1.06-1.36), the aHR for HSV-2 was 1.03 (0.93-1.14), and the aHR for CMV was 1.35 (1.23-1.48) (**Figure 1** and **Table 3**). When stratified by cohort, the aHR for HSV-1 was 1.31 (1.12-1.52) in the early cohort and 1.00 (0.82-1.23) in the late cohort (**Figure 2** and **Table 3**). For HSV-2, the aHRs were 0.95 (0.84-1.08) and 1.19 (1.02-1.39) for the early and late cohorts, respectively, and for CMV the aHRs were 1.44 (1.28-1.62) and 1.19 (1.02-1.40), respectively (**Figure 2** and **Table 3**). The results for univariate analyses for our selection of variables chosen *a priori* are included in **Supplemental Table 3.**

**Figure 1:**
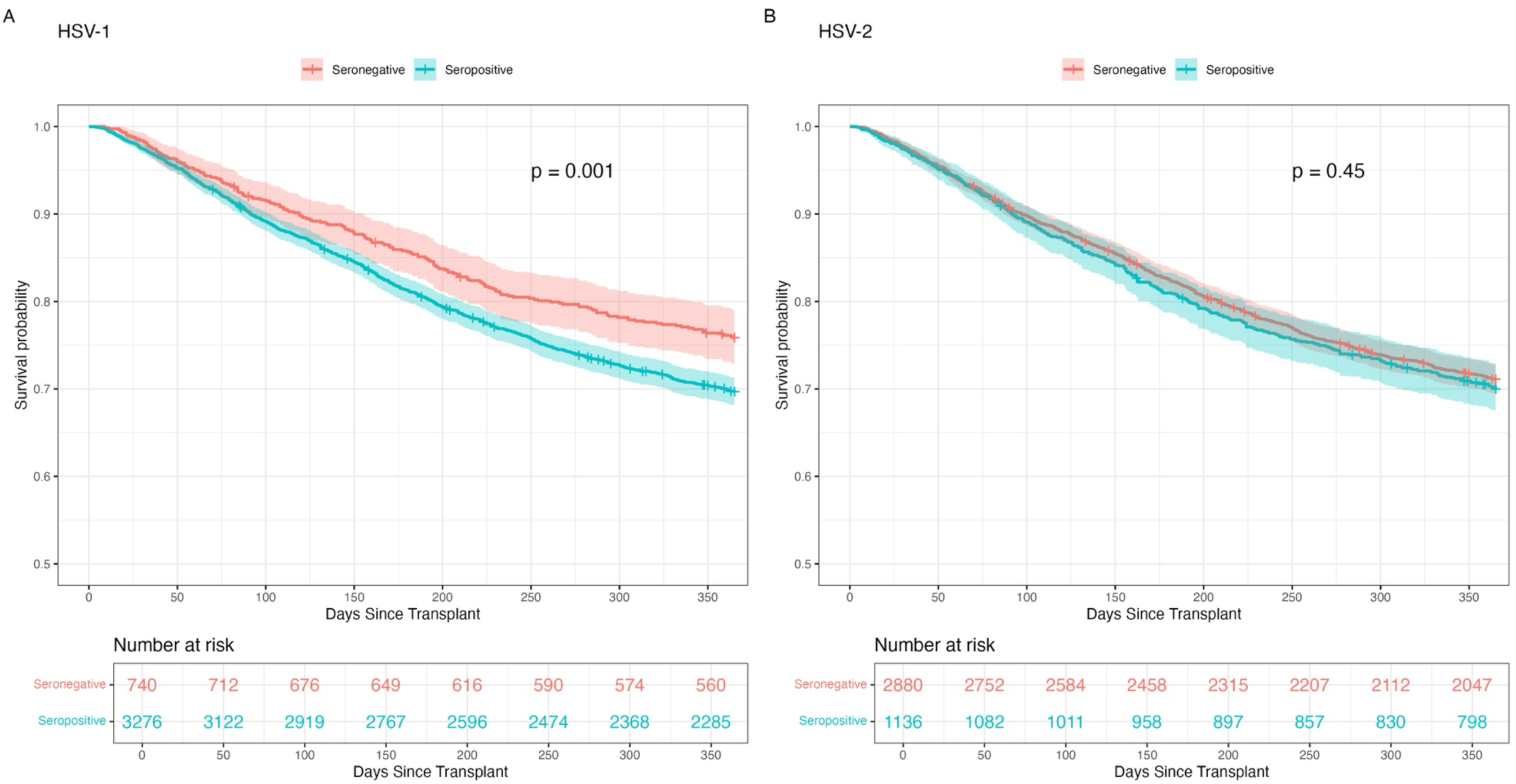
Kaplan-Meier curves comparing 1-year survival between (A) HSV-1 and (B) HSV-2 seropositive and seronegative patients, 2002-2022.

**Table 3:** Adjusted hazard ratios for 1-year mortality comparing HSV and CMV seropositive to seronegative patients overall and by cohort.

|  | Overall (2002-2022) |  | Early (2002-2012) |  | Late (2013-2022) |  |
| --- | --- | --- | --- | --- | --- | --- |
|  | aHR | 95% CI | aHR | 95% CI | aHR | 95% CI |
| **HSV-1** | 1.20 | (1.06-1.36) | 1.31 | (1.12-1.52) | 1.00 | (0.82-1.23) |
| **HSV-2** | 1.03 | (0.93-1.14) | 0.95 | (0.84-1.08) | 1.19 | (1.02-1.39) |
| **CMV** | 1.35 | (1.23-1.48) | 1.44 | (1.28-1.62) | 1.19 | (1.02-1.40) |

*HSV models adjusted for age, sex, conditioning intensity, underlying disease risk, HLA matching and graft type, CMV serostatus, aGVHD (time-varying), and year of transplant. CMV model adjusted for HSV-1 serostatus in lieu of CMV serostatus.*

**Figure 2:**
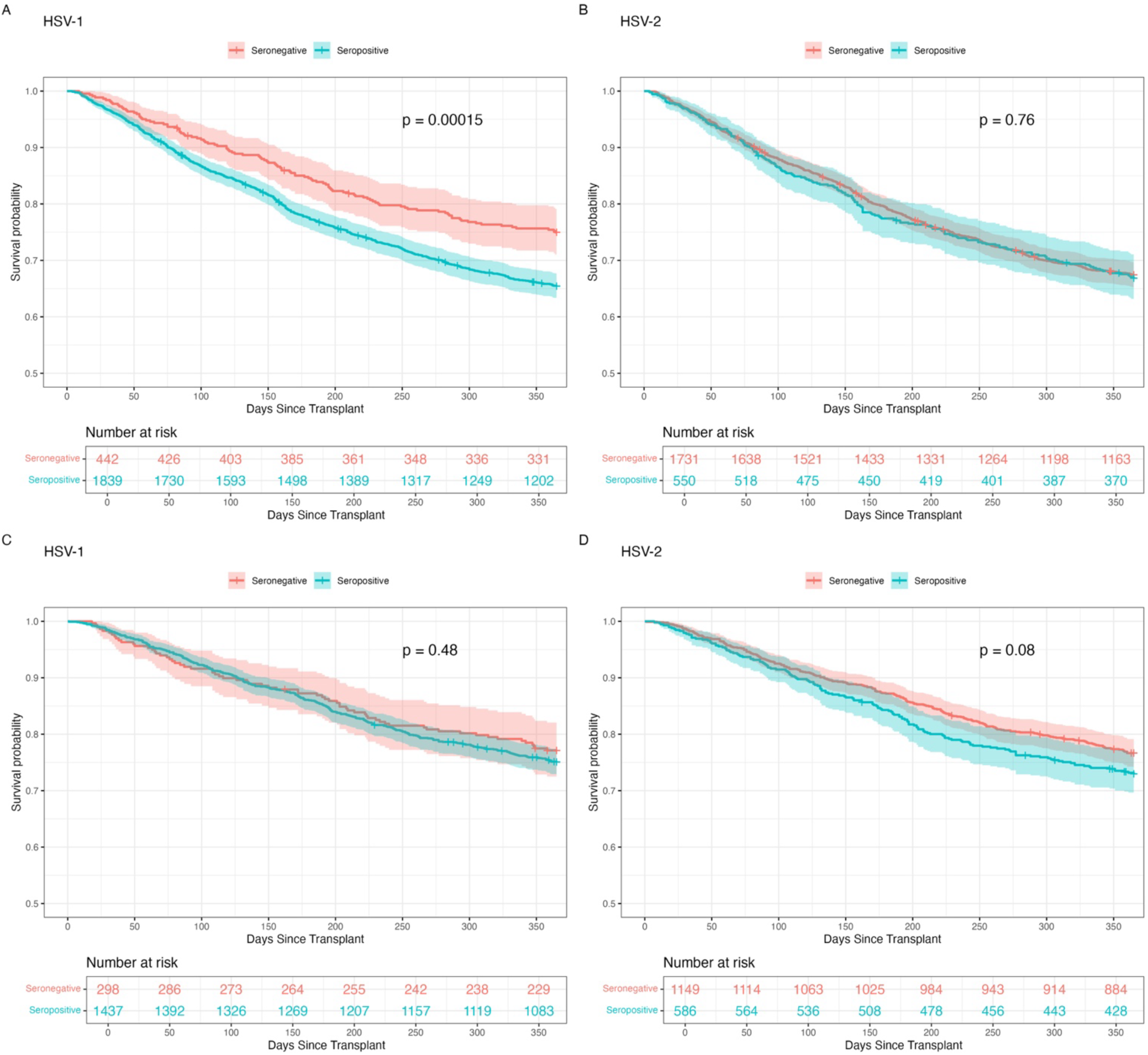
Kaplan-Meier curves comparing 1-year survival between HSV-1 and HSV-2 seropositive and seronegative patients by cohort: Early 2002-2012 (A-B) and Late 2013-2022 (C-D)

### HSV-1, HSV-2, and CMV Seropositivity and Relapse, Relapse-Related Mortality (RRM), and Non-Relapse Mortality (NRM)

After excluding 134 (3.3%) patients with nonmalignant conditions, aplastic anemia or solid tumors, a total of 3,882 patients were included in the analyses of relapse, RRM, and NRM. Unadjusted cumulative incidence curves for relapse, RRM, and NRM by HSV-1 by HSV-1/2 serostatus are shown in **Supplemental Figures 2-4** for the different cohorts (early, late, and overall), and the adjusted hazard ratios are included in **Table 4**.

**Table 4:** Adjusted overall (2002-2022) hazard ratios for relapse, RRM, and NRM.

|  | NRM |  | RRM |  | Relapse |  |
| --- | --- | --- | --- | --- | --- | --- |
|  | aHR | 95% CI | aHR | 95% CI | aHR | 95% CI |
| **HSV-1** | 1.04 | (0.89-1.21) | 1.75 | (1.40-2.19) | 1.46 | (1.24-1.71) |
| **HSV-2** | 0.99 | (0.87-1.13) | 1.13 | (0.97-1.33) | 1.16 | (1.03-1.31) |
| **CMV** | 1.35 | (1.19-1.53) | 1.35 | (1.16-1.58) | 1.06 | (0.94-1.19) |

*NRM = non-relapse mortality, RRM = relapse-related mortality*
- 1. NRM Model: Primary Model; endpoint of death not preceded by relapse, observations censored at time of relapse for patients who relapsed within 1 year* - 2. RRM Model: Primary Model; endpoint of death preceded by relapse, observations censored at time of death for patients who did not relapse within 1 year* - 3. Relapse Model: Primary Model; endpoint of relapse, observations censored at time of death for patients who did not relapse within 1 year*

#### HSV-1

For HSV-1 the overall aHRs for relapse, RRM, and NRM were 1.46 (1.24-1.71), 1.75 (1.40-2.19) and 1.04 (0.89-1.21), respectively. The aHR for relapse in the early cohort was 1.54 (1.25-1.90) versus 1.28 (1.00-1.64) in the late cohort. The aHR for RRM in the early cohort was 1.95 (1.46-2.61) compared to 1.39 (0.97-1.99) in the late cohort. The aHR for NRM was 1.17 (0.97-1.41) in the early cohort and 0.83 (0.65-1.07) in the late cohort.

#### HSV-2

For HSV-2, the overall aHRs for relapse, RRM, and NRM were 1.16 (1.03-1.31) 1.13 (0.97-1.33) and 0.99 (0.87-1.13), respectively. The aHR for relapse was 1.11 (0.94-1.32) in the early cohort and 1.22 (1.02-1.47) in the late cohort. The aHR for RRM was 0.93 (0.75-1.15) in the early cohort and 1.58 (1.24-2.03) in the late cohort. The aHR for NRM was 1.00 (0.85-1.18) in the early cohort and 1.00 (0.81-1.24) in the late cohort.

#### CMV

For CMV, the overall aHRs for relapse, RRM, and NRM were 1.06 (0.94-1.19), 1.35 (1.16-1.58), and 1.35 (1.19-1.53), respectively. The aHR for relapse was 1.18 (1.01-1.37) in the early cohort and 0.94 (0.79-1.13) in the late cohort. The aHR for RRM was 1.68 (1.38-2.04) in the early cohort and 0.98 (0.76-1.26) in the late cohort. The aHR for NRM was 1.37 (1.18-1.59) in the early cohort and 1.30 (1.05-1.60) in the late cohort.

### Sensitivity Analyses

#### HSV and Mortality

Adjusted hazard ratios from the sensitivity analyses are included in **Supplemental Table 4**. From the overall models, adjusting for time-varying CMV reactivation did not meaningfully change the hazard ratios for HSV-1 (aHR: 1.22 vs. 1.20) or HSV-2 (aHR: 1.01 vs. 1.03). Removing aGVHD from the model resulted in a slight increase in the point estimate for the hazard for HSV-1 (aHR: 1.26 vs. 1.20) but little change for HSV-2 (aHR: 1.00 vs 1.03) or CMV (aHR: 1.37 vs 1.35). Reclassifying 341 indeterminate HSV results as seronegative also did not meaningfully change any hazard ratios (**Supplemental Table 4**). We did not find evidence in the overall model to conclude that CMV serostatus or the conditioning regimen modified the association between HSV-1 or HSV-2 and mortality (**Supplemental Tables 5 and 6**).

#### HSV, CMV, and Blood Transfusions

We noted a higher HSV seroprevalence in the FHCC aHCT population than in the US general population across age strata (**Table 2**). To explore whether blood transfusions, which may be associated both with higher risk disease and possible passive transfer of HSV and CMV antibodies, were associated with seropositivity for these viruses, we conducted an exploratory analysis to evaluate the association between the number of pre-WB blood transfusions and HSV and CMV serostatus among a subgroup of more recent transplant recipients. Among 449 patients who had a first aHCT between 2020-2022 and a valid HSV WB, 228 (50.8%) had at least one red blood cell (RBC) transfusion-related code linked with their records within 90 days of HSV WB. Among these 228 patients, the median number of pre-HSV WB RBC transfusions was 8 (IQR: 4-14). In both univariate and multivariable models, RBC transfusions were associated with higher odds of both HSV-1, HSV-2, and CMV seropositivity. A one-unit increase in the number of RBC transfusions was associated with 7% higher odds of HSV-1 seropositivity (OR: 1.07 [1.02-1.12]), 4% higher odds of HSV-2 seropositivity (OR: 1.04 [1.01-1.07]), and 4% higher odds of CMV seropositivity (OR: 1.04 [1.01-1.07]. When adjusting for age and underlying disease risk, which may be associated with both likelihood of seropositivity and number of transfusions, the estimates were generally unchanged (data not shown).

## Discussion

In this large cohort of aHCT recipients in the era of universal antiviral prophylaxis, we found that HSV-1 and CMV seropositive patients had significantly higher hazards of all-cause mortality during the first year post-transplant compared to seronegative patients, while HSV-2 seropositivity was overall not associated with mortality. Notably, these associations were observed after adjusting for major transplant-related risk factors for mortality, including recipient CMV reactivation post-transplant, disease risk, conditioning regimen intensity, and aGVHD. The associations between HSV-1 and all-cause mortality waned over time, while the association between HSV-2 and relapse/RRM appeared stronger.

The overall risk of mortality declined over the study period. As described by McDonald et al., numerous incremental changes to transplant practices over the past few decades have likely contributed to the overall improvement in survival between the early and late cohorts.^1^ However, understanding the specific mechanisms that may underlie the change in the magnitude of the HSV and CMV hazard ratios over time is challenging. The HSV/VZV prophylaxis strategy was unchanged at FHCC over the study period and is thus unlikely to explain this difference; we did not assess the impact of letermovir prophylaxis on mortality between CMV-seropositive and seronegative recipients. The availability of the Armand DRI score^31^ for patients who transplanted in 2013 and later may have resulted in better adjustment for confounding by underlying disease risk in the later cohort, but is also unlikely to fully account for this change. The overall reduction in the number of deaths over time coupled with the higher HSV-1 seroprevalence also results in lower statistical power of the model in the later cohort and less stable estimates, which further complicates interpretation of these trends over time.

Contrary to our hypotheses, the overall survival difference between HSV seropositive and seronegative patients was driven by differences in relapse and RRM rather than NRM. HSV-1 was overall associated with higher hazards of relapse and RRM, although the strength of this association waned over time. CMV was associated with relapse, RRM, and NRM in the early cohort, but only appeared to be associated with NRM in the late cohort. HSV-2 was overall significantly associated with relapse, driven by the association in the later cohort. CMV seropositivity was consistently associated with a higher hazard of non-relapse mortality over time, suggesting that the observed associations between HSV and mortality are unlikely to be fully explained by potential residual confounding by CMV.

The proportion of HSV-seropositive patients in this cohort was substantially higher than what has been reported in national seroprevalence estimates from the National Health and Nutrition Examination Survey, particularly among younger age groups.^32^ While HSV seroprevalence data among adult aHCT recipients at other US centers are lacking, two studies conducted among pediatric aHCT recipients have reported a seroprevalence over 50% in this population.^33,34^ Prior studies from FHCC that included aHCT patients from 1992-2003 have also demonstrated high (>70%) HSV seropositivity that appears to be increasing over time,^12,27^ in contrast to national trends.^32^ Given these observations, we were concerned about the possibility of passive HSV antibody transfer during pre-transplant blood transfusions that could result in a positive HSV WB result. Evidence suggests that passive CMV IgG transfer can occur from pre-transplant supportive red blood cell and/or platelet transfusions (including those that have undergone leukodepletion), and lead to misclassification of the recipient’s serostatus.^35–37^ We found a positive association between pre-transplant RBC transfusions and odds of HSV and CMV seropositivity, suggesting that passive transfer of HSV antibodies may also be possible. Furthermore, prior research suggests that transfusions may be associated with increased mortality among patients with various hematologic malignancies,^38–41^ although the potential mechanisms underlying this association are unclear. As a result, it is possible that the observed association between HSV seropositivity and relapse/relapse-related mortality is confounded by transfusion history. Additional research is needed to establish proof-of-principle of HSV antibody transfer and to further examine the relationship between RBC and/or platelet transfusions and HSV/CMV serostatus.

Data on the association between HSV serostatus and post-aHCT survival in the current era of long-term prophylaxis are sparse. A prior study conducted at FHCC among aHCT patients with leukemia and HLA-matched sibling donors between 1992 and 2003 found that among high-risk patients, HSV seropositivity was associated with higher hazards of relapse (aHR = 1.9 [1.2-3.0]) and mortality (aHR = 2.2 [1.5-3.3]), adjusting for similar factors including CMV serostatus.^42^ HSV seropositive patients in this study received ACV prophylaxis for only the first 30 days until the year 2000, when the duration of prophylaxis was extended to 1 year post-transplant for VZV seropositive patients,^42^ and thus most patients did not receive long-term ACV.

Other studies have explored the potential relationship between HSV-1 shedding/reactivation and mortality despite prophylaxis with mixed findings. For example, with respect to oral HSV shedding, Guimaraes et al. found that aHCT patients with shedding prior to transplant (and prior to ACV prophylaxis) had significantly worse survival than those who did not, while there was no survival difference among those who did vs. did not shed after HCT who survived to at least day 100.^16^ Miranda-Silva et al. observed that oral HSV-1 shedding at nadir was associated with a higher risk of relapse,^43^ and Kakiuchi et al. reported that patients with oral HSV-1 shedding had a significantly higher (HR: 2.44; 95% CI = 1.35–4.39) hazard for mortality within 100 days than patients without, independent of clinical HSV disease.^18^ With respect to viremia, Tang et al. found no statistically significant differences between patients with HSV viremia and those without with respect to overall survival, disease-free survival, or NRM^44^, and Przybylski et al. also found no significant difference in the cumulative incidence of 100-day or 180-day mortality between patients with viremia and those without.^45^ These studies all used varying sample types, testing frequencies, and prophylaxis strategies, which may contribute to some of the heterogeneity in findings. Overall, these results highlight the need for future prospective studies with routine HSV-1 monitoring to better understand whether HSV serostatus and viral shedding, either directly or as an indirect marker of another risk factor, could be predictive of adverse post-transplant outcomes.

### Strengths and Limitations

This study has several key strengths. Pre-transplant HSV serologic testing using the gold-standard UW WB assay for all aHCT patients at FHCC provided a unique opportunity to analyze the HSV-mortality association in a cohort of patients with robust exposure measurement. The routine use of long-term, high-dose ACV/VACV prophylaxis for VZV/HSV prevention in this population throughout the first year post transplant also helps minimize the possibility of confounding due to differences in prophylaxis dose or duration. However, because daily antiviral use (particularly during outpatient care) is not routinely assessed, patient adherence to antiviral prophylaxis regimens was assumed rather than measured directly.

This analysis also has a few important limitations. First, in the later cohort, our study may have been underpowered to detect differences in mortality associated with HSV-1 and CMV due to several factors, including a higher seroprevalence of both viruses, a lower number of transplants performed in this cohort (due to fewer years of data and a fewer transplants performed per year), and a lower cumulative mortality, resulting in a smaller number of events. Second, despite adjusting for several risk factors for mortality, there are additional known factors (e.g. socioeconomic status) associated with both HSV serostatus and mortality, and failure to adjust for these may have biased our estimates. Our data also come from a single US transplant center, and therefore our findings may not be generalizable to other centers. The exclusive use of the UW WB assay also makes generalizability to other centers challenging. Finally, HSV serostatus of the donor is not routinely assessed at FHCC and, therefore, could not be incorporated into these analyses.

### Summary and Future Directions

We have shown that despite antiviral prophylaxis, HSV-1 seropositivity is associated with one-year mortality among aHCT recipients at a single US transplant center and is specifically associated with higher hazards of relapse and relapse-related mortality. These associations waned over time. CMV seropositivity was also associated with higher one-year relapse-related and non-relapse mortality. HSV-2 serostatus was overall not associated with mortality. Our preliminary findings suggest that CMV and HSV serostatus may be associated with pre-transplant blood transfusion history. Overall, these results highlight the need for further research to evaluate whether the observed relationship between HSV serostatus, relapse and mortality suggests that HSV may be causally implicated in adverse post-transplant survival outcomes.

## Data Sharing Statement

Data may be made available upon reasonable request from the corresponding author.

## Acknowledgements

This work was supported by the National Cancer Institute (NCI-P30CA0087-48). This work has been submitted in partial fulfillment of the requirement for a PhD for MDF. We would like to thank Chris Davis for data curation, management and abstraction, and Zach Stednick for work on the preliminary analysis. We also thank all patients who contributed their data for scientific research, and are grateful for the healthcare providers, pharmacists, social workers, and support staff who cared for them.

## Authorship Contributions

### Contributions

SAP, CJ, MB and TG conceptualized the study; all authors contributed to methodology; CJ, MJB, MAB, BMS, SAP, AW, DJM ESF conducted the investigation; MDF analyzed results, created figures, and wrote the original draft; SAP, CJ, AIP, and RLW provided supervision; all authors reviewed and edited final manuscript.

## SUPPLEMENTAL MATERIALS

**Supplemental Table 1:** Description of categorical covariates.

| Variable | Levels | Description |
| --- | --- | --- |
| **Donor matching and graft type** | 1 | Mismatched Sibling, Matched Non-Sibling, or Mismatched Non-Sibling; PBSC* |
|  | 2 | Matched Unrelated; PBSC |
|  | 3 | Unrelated Mismatched, PBSC |
|  | 4 | Cord Blood (Mismatched) |
|  | 5 | Matched Unrelated, BM<sup>2</sup> |
|  | 6 | Mismatched Sibling, Matched Non-Sibling, or Mismatched Non-Sibling; BM |
|  | 7 | Unrelated Mismatched; BM |
|  | 8 | Matched Sibling; BM |
|  | 9 | Matched Sibling; PBSC |
| **2002-2012: Disease Risk<sup>1,24</sup> \*\*** | Low | *Examples:* chronic myeloid leukemia (chronic phase or remission), paroxysmal nocturnal hemoglobinuria, aplastic anemia (relapse or remission), systemic sclerosis, chronic granulomatous disease, pure red cell anemia, and immune diseases, disorders or syndromes |
|  | Intermediate | *Examples:* acute leukemia or lymphoma (in remission), chronic myeloid leukemia (accelerated phase, in remission following blast crisis), multiple myeloma (in remission), chronic lymphocytic leukemia (in remission), non-Hodgkin's lymphoma (in remission), Hodgkin's disease (in remission), some myelodysplastic syndromes |
|  | High | *Examples:* chronic myeloid leukemia (in blast crisis), acute leukemia or lymphoma in relapse, non-Hodgkin's lymphoma (in relapse), Hodgkin's disease (in relapse), solid tumors, some myelodysplastic syndromes |
|  | Unknown | Various conditions with unknown or missing status (*Example:* chronic myeloid leukemia, status unknown) |
| **2013-2022: Armand Disease Risk Index (DRI)<sup>31</sup> \*\*\*** | Low | *Examples:* various diseases in complete remission (AML w/ favorable cytogenetics, CLL, HL, indolent NHL) or partial remission (CLL, indolent NHL), CML in 1<sup>st</sup> or 2<sup>nd</sup> chronic phase |
|  | Intermediate | *Examples:* CML in advanced phase, AML in complete remission w/ intermediate cytogenetics, MM, low-risk MDS w/ adverse cytogenetics, HL in partial remission, advanced indolent NHL |
|  | High | *Examples:* advanced HL, high-risk early stage MDS with adverse cytogenetics, advanced AML with intermediate cytogenetics, |
|  | Very High | *Examples:* CML in blast phase, advanced aggressive NHL, advanced ALL, advanced AML with adverse cytogenetics |
|  | Unknown | Various conditions with unknown or missing status<br>(*Examples:* missing or TBD cytogenetics, TBD cytogenetics, missing disease status, disease not classifiable) |
| **Conditioning intensity\*\*\*\*** | High | *Examples:* BU + CY<br>CY + LI(400) + H-TBI(1200)<br>TREO + FLU + TBI(200) |
|  | Intermediate | *Examples:* CY + FLU + TBI(200)<br>CY + H-TBI(1200)<br>L-PAM + FLU |
|  | Low | *Examples:* FLU + TBI(200)<br>FLU + TBI(300)<br>TBI (200) |

\* PBSC = peripheral blood stem cells; BM = bone marrow
\*\* Disease severity as described by Gooley et al. and McDonald et al. based on expert review
\*\*\* The Armand DRI score was available for patients who received a transplant in 2013 and later. In the full model, the scores were combined into a composite disease risk score: we used the 2002-2012 scores for the early (2002-2012) cohort and DRI for the later (2013-2022) cohort. For patients who had transplants in the later cohort and had missing DRI scores (ex: due to missing cytogenetics data), the 2002-2012 grading system was used.
\*\*\*\*Conditioning intensity was based on the TCI score combined with good clinical judgment from an expert oncologist (Melinda A. Biernacki). Three examples of regimens are provided above for each intensity level. BU = busulfan, CY = cyclophosphamide, FLU = fludarabine, TBI = total body irradiation, TREO = treosulfan, L-PAM = melphalan

**Supplemental Table 2:** Underlying disease categorization.

| **Overall Category** | **Conditions** |
| --- | --- |
| Acute Lymphoblastic Leukemia (ALL) | Acute lymphoblastic leukemia |
| Acute Myeloid Leukemia (AML) | Erythroleukemia, acute leukemia NOS, acute megakaryoblastic leukemia, acute myeloid leukemia, acute myelomonocytic leukemia, acute monocytic leukemia, acute promyelocytic leukemia, biphenotypic leukemia (AML vs ALL), leukemia NOS, myeloid leukemia NOS, mast cell leukemia |
| Aplastic Anemia/ Paroxysmal Nocturnal Hemoglobinuria (PNH) | Aplastic anemia NOS, aplastic anemia (constitutional Fanconi's anemia), paroxysmal nocturnal hemoglobinuria |
| Chronic Lymphocytic Leukemia (CLL) | Chronic lymphocytic leukemia |
| Chronic Myeloid Leukemia (CML) | Chronic myeloid leukemia |
| Hodgkin's Lymphoma | Mixed cellularity HD NOS, lymphocytic predominant HD NOS, nodular sclerosis HD NOS, HD NOS |
| Myelodysplastic Syndromes and Myeloproliferative Neoplasms (MDS/MPN) | Blastic plasmacytoid dendritic cell neoplasm, chronic leukemia NOS, chronic myelomonocytic leukemia, eosinophilic leukemia, fibrosis NOS (myelofibrosis), myelodysplastic syndrome (preleukemia), myeloid (agnogenic) metaplasia, myeloproliferative disease NOS, malignant neoplasm, polycythemia vera, refractory anemia NOS, refractory anemia with excess blasts (RAEB), RAEB "in transformation", sideroblastic anemia acquired, refractory cytopenia with multilineage dysplasia, thrombocythemia, thrombocytosis |
| Lymphoma (Non-Hodgkin's) (NHL) | Burkitt's lymphoma NOS, non-Hodgkin's malignant lymphoma NOS, follicular malignant lymphoma NOS, immunoblastic malignant lymphoma, large cell diffuse malignant lymphoma, large cell follicular malignant lymphoma NOS, lymphoblastic malignant lymphoma, lymphoplasmacytoid type malignant lymphoma, Mantle Cell malignant lymphoma, mixed small cleaved large cell malignant lymphoma, malignant lymphoma NOS, small cleaved cell follicular malignant lymphoma, small lymphocytic malignant lymphoma, Waldenstrom's macroglobulinemia |
| Myeloma (M) | Multiple myeloma, monoclonal gammopathy, plasma cell leukemia |
| Nonmalignant (NM) | Chronic granulomatous disease, congenital dyserythropoietic anemia, Crohn's disease NOS, dyskeratosis congenita, eosinophilic Syndrome NOS, erythropoietic protophyria, familial hemophagocytic histiocytosis, hemoglobin S disease (sickle cell), hemophagocytic lymphohistiocytosis, IgA nephropathy (Berger disease), immune deficiency disorder/disease/syndrome, mastocytoma NOS, multiple sclerosis, progressive systemic sclerosis, thalassemic syndrome NOS, chronic pure red cell anemia |
| Other Malignant (OM) | Colon carcinoma, hairy cell leukemia, mycosis fungoides, renal cell carcinoma, prolymphocytic leukemia, sarcoma NOS |
| Plasma Cell Neoplasms | Primary amyloidosis, multiple myeloma, monoclonal gammopathy,<br>plasma cell leukemia |

\* *NOS* = *not otherwise specified*

**Supplemental Table 3:** Univariate Cox proportional hazards model results for 1-year overall mortality.

|  |  | Full Cohort (2002-2022) |  | Early Cohort (2002-2012) |  | Late Cohort (2013-2022) |  |
| --- | --- | --- | --- | --- | --- | --- | --- |
| Variable | Levels | HR (95% CI) | p-value | HR (95% CI) | p-value | HR (95% CI) | p-value |
| HSV-1 Serostatus | Seronegative | (Ref) |  | (Ref) |  | (Ref) |  |
|  | Seropositive | 1.31 (1.11-1.53) | <0.01 | 1.46 (1.20-1.79) | <0.01 | 1.11 (0.86-1.44) | 0.43 |
| HSV-2 Serostatus | Seronegative | (Ref) |  | (Ref) |  | (Ref) |  |
|  | Seropositive | 1.04 (0.92-1.18) | 0.55 | 1.02 (0.86-1.20) | 0.85 | 1.19 (0.97-1.44) | 0.09 |
| Age | Continuous (years) | 1.01 (1.00-1.01) | <0.01 | 1.01 (1.01-1.02) | <0.01 | 1.01 (1.01-1.02) | <0.01 |
| Sex | Female | (Ref) |  | (Ref) |  | (Ref) |  |
|  | Male | 1.16 (1.03-1.31) | 0.01 | 1.16 (1.00-1.34) | 0.04 | 1.15 (0.95-1.39) | 0.16 |
| Conditioning Intensity | Low | (Ref) |  | (Ref) |  |  |  |
|  | Intermediate | 0.99 (0.82-1.18) | 0.88 | 1.65 (1.26-2.15) | <0.01 | 0.83 (0.63-1.08) | 0.16 |
|  | High | 1.10 (0.96-1.25) | 0.18 | 1.23 (1.05-1.45) | 0.01 | 0.89 (0.71-1.13) | 0.34 |
| Underlying Disease Risk | Low | (Ref) |  | (Ref) |  | (Ref) |  |
|  | Intermediate | 1.38 (1.06-1.78) | 0.02 | 1.33 (0.93-1.91) | 0.12 | 1.31 (0.90-1.90) | 0.16 |
|  | High | 2.40 (1.85-3.13) | <0.01 | 2.07 (1.44-2.97) | <0.01 | 2.53 (1.71-3.73) | <0.01 |
|  | Very High | 3.78 (2.39-5.97) | <0.01 | N/A |  | 4.56 (2.71-7.68) | <0.01 |
|  | Unknown | 1.31 (0.74-2.35) | 0.36 | 1.22 (0.63-2.36) | 0.58 | 0.92 (0.22-3.86) | 0.91 |
| Recipient CMV Serostatus | Seronegative | (Ref) |  | (Ref) |  | (Ref) |  |
|  | Seropositive | 1.40 (1.24-1.58) | <0.01 | 1.57 (1.35-1.83) | <0.01 | 1.19 (0.97-1.45) | 0.09 |
|  | Missing | 1.48 (0.87-2.52) | 0.15 | 1.57 (0.65-3.81) | 0.32 | 1.57 (0.80-3.07) | 0.19 |
| HLA Matching and Graft Type | Cord Blood (Mismatched) | (Ref) |  | (Ref) |  | (Ref) | <0.005 |
|  | Mismatched Sibling, Matched Non-Sib., or Mismatched Non-Sib.; PBSC | 0.76 (0.54-1.08) | 0.12 | 0.49 (0.28-0.88) | 0.02 | 1.02 (0.66-1.59) | 0.92 |
|  | Matched Unrelated; PBSC | 0.65 (0.53-0.81) | <0.01 | 0.64 (0.47-0.86) | <0.01 | 0.61 (0.45-0.83) | <0.01 |
|  | Unrelated Mismatched, PBSC | 1.06 (0.83-1.36) | 0.62 | 1.01 (0.73-1.41) | 0.95 | 0.93 (0.64-1.35) | 0.70 |
|  | Matched Unrelated, BM | 0.61 (0.43-0.85) | <0.01 | 0.56 (0.37-0.84) | <0.01 | 0.38 (0.17-0.84) | 0.02 |
|  | Mismatched Sib., Matched Non-Sib., or Mismatched Non-Sib.; BM | 1.23 (0.86-1.75) | 0.26 | 1.00(0.65-1.53) | 0.98 | 1.24 (0.61-2.52) | 0.54 |
|  | Unrelated Mismatched; BM | 1.33 (0.88-2.00) | 0.18 | 1.05 (0.66-1.67) | 0.84 | 1.14 (0.28-4.70) | 0.85 |
|  | Matched Sib.; BM | 0.57 (0.33-0.97) | 0.04 | 0.66 (0.36-1.19) | 0.17 | 0.12 (0.02-0.87) | 0.04 |
|  | Matched Sib.; PBSC | 0.66 (0.53-0.83) | <0.01 | 0.53 (0.39-0.72) | <0.01 | 0.75 (0.54-1.05) | 0.10 |
| aGVHD | No | *(Ref)* |  | *(Ref)* |  | *(Ref)* |  |
|  | Yes (indicator: grade 2 or higher) | 0.99 (0.87-1.11) | 0.81 | 1.01 (0.87-1.18) | 0.88 | 0.90 (0.74-1.09) | 0.29 |
| Year of Transplant | Continuous (years) | 0.96 (0.95-0.97) | <0.01 | 0.96 (0.94-0.98) | <0.01 | 0.93 (0.90-0.96) | <0.01 |

**Supplemental Table 4:**
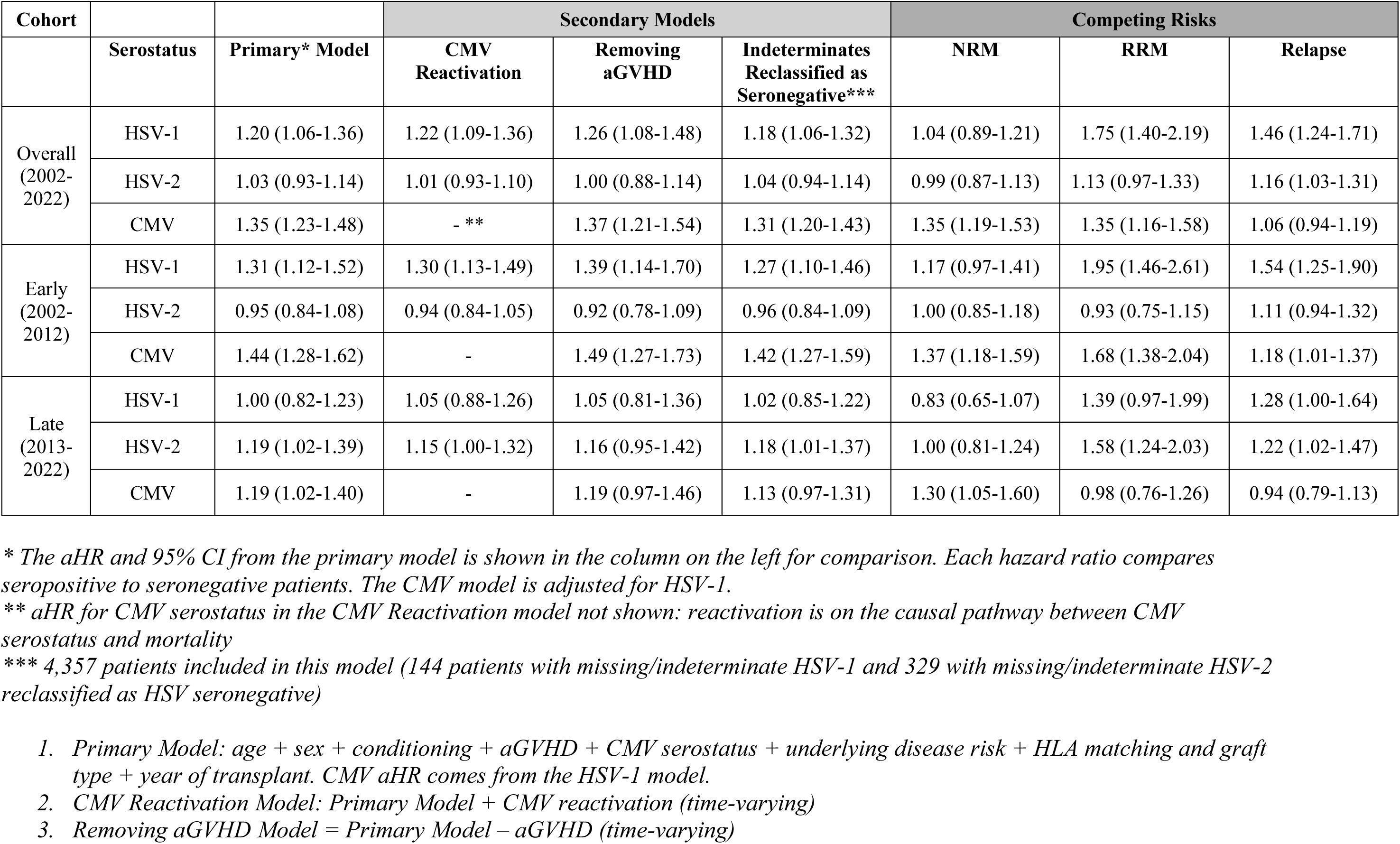
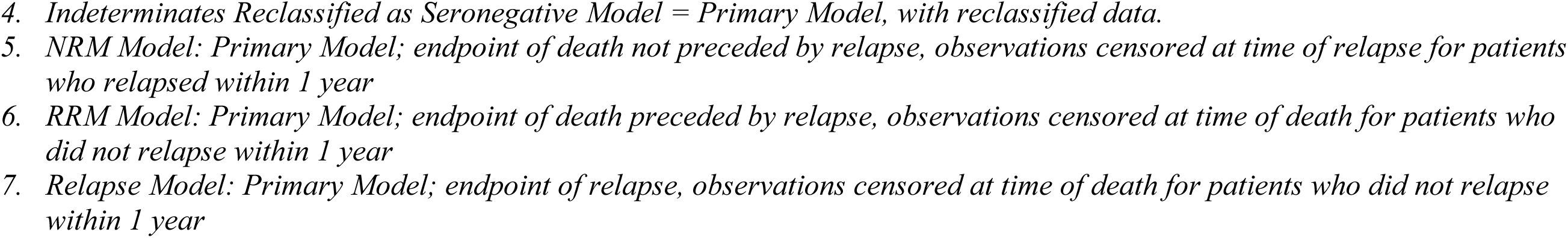
Adjusted hazard ratios and 95% confidence intervals for 1-year mortality associated with HSV-1, HSV-2, and CMV from sensitivity analyses.

| Cohort |  |  | Secondary Models |  |  | Competing Risks |  |  |
| --- | --- | --- | --- | --- | --- | --- | --- | --- |
|  | Serostatus | Primary* Model | CMV<br>Reactivation | Removing<br>aGVHD | Indeterminates<br>Reclassified as<br>Seronegative*** | NRM | RRM | Relapse |
| Overall<br>(2002-2022) | HSV-1 | 1.20 (1.06-1.36) | 1.22 (1.09-1.36) | 1.26 (1.08-1.48) | 1.18 (1.06-1.32) | 1.04 (0.89-1.21) | 1.75 (1.40-2.19) | 1.46 (1.24-1.71) |
|  | HSV-2 | 1.03 (0.93-1.14) | 1.01 (0.93-1.10) | 1.00 (0.88-1.14) | 1.04 (0.94-1.14) | 0.99 (0.87-1.13) | 1.13 (0.97-1.33) | 1.16 (1.03-1.31) |
|  | CMV | 1.35 (1.23-1.48) | - ** | 1.37 (1.21-1.54) | 1.31 (1.20-1.43) | 1.35 (1.19-1.53) | 1.35 (1.16-1.58) | 1.06 (0.94-1.19) |
| Early<br>(2002-2012) | HSV-1 | 1.31 (1.12-1.52) | 1.30 (1.13-1.49) | 1.39 (1.14-1.70) | 1.27 (1.10-1.46) | 1.17 (0.97-1.41) | 1.95 (1.46-2.61) | 1.54 (1.25-1.90) |
|  | HSV-2 | 0.95 (0.84-1.08) | 0.94 (0.84-1.05) | 0.92 (0.78-1.09) | 0.96 (0.84-1.09) | 1.00 (0.85-1.18) | 0.93 (0.75-1.15) | 1.11 (0.94-1.32) |
|  | CMV | 1.44 (1.28-1.62) | - | 1.49 (1.27-1.73) | 1.42 (1.27-1.59) | 1.37 (1.18-1.59) | 1.68 (1.38-2.04) | 1.18 (1.01-1.37) |
| Late<br>(2013-2022) | HSV-1 | 1.00 (0.82-1.23) | 1.05 (0.88-1.26) | 1.05 (0.81-1.36) | 1.02 (0.85-1.22) | 0.83 (0.65-1.07) | 1.39 (0.97-1.99) | 1.28 (1.00-1.64) |
|  | HSV-2 | 1.19 (1.02-1.39) | 1.15 (1.00-1.32) | 1.16 (0.95-1.42) | 1.18 (1.01-1.37) | 1.00 (0.81-1.24) | 1.58 (1.24-2.03) | 1.22 (1.02-1.47) |
|  | CMV | 1.19 (1.02-1.40) | - | 1.19 (0.97-1.46) | 1.13 (0.97-1.31) | 1.30 (1.05-1.60) | 0.98 (0.76-1.26) | 0.94 (0.79-1.13) |

\* The aHR and 95% CI from the primary model is shown in the column on the left for comparison. Each hazard ratio compares seropositive to seronegative patients. The CMV model is adjusted for HSV-1.
\*\* aHR for CMV serostatus in the CMV Reactivation model not shown: reactivation is on the causal pathway between CMV serostatus and mortality
\*\*\* 4,357 patients included in this model (144 patients with missing/indeterminate HSV-1 and 329 with missing/indeterminate HSV-2 reclassified as HSV seronegative)
1. Primary Model: age + sex + conditioning + aGVHD + CMV serostatus + underlying disease risk + HLA matching and graft type + year of transplant. CMV aHR comes from the HSV-1 model. 2. CMV Reactivation Model: Primary Model + CMV reactivation (time-varying) 3. Removing aGVHD Model = Primary Model – aGVHD (time-varying)
4. *Indeterminates Reclassified as Seronegative Model = Primary Model, with reclassified data.* 5. *NRM Model: Primary Model; endpoint of death not preceded by relapse, observations censored at time of relapse for patients who relapsed within 1 year* 6. *RRM Model: Primary Model; endpoint of death preceded by relapse, observations censored at time of death for patients who did not relapse within 1 year* 7. *Relapse Model: Primary Model; endpoint of relapse, observations censored at time of death for patients who did not relapse within 1 year*

**Supplemental Table 5:**
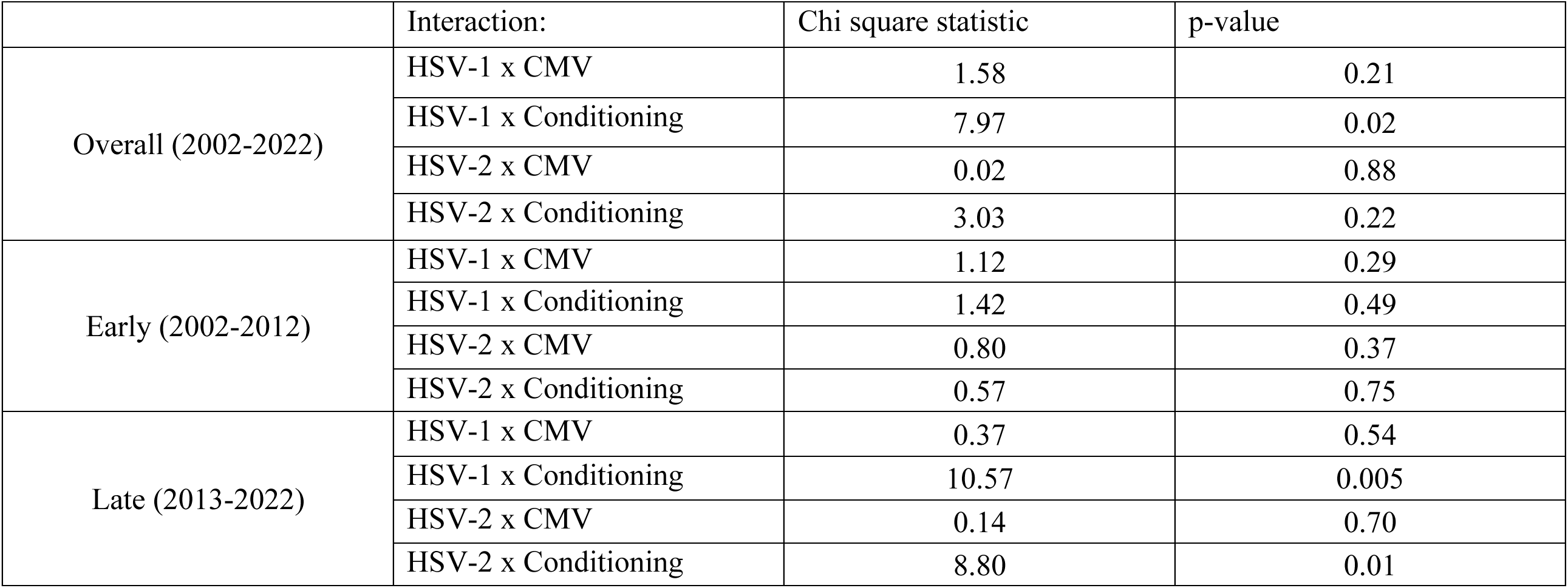
Model ANOVA results for interaction.

**Supplemental Table 6:**
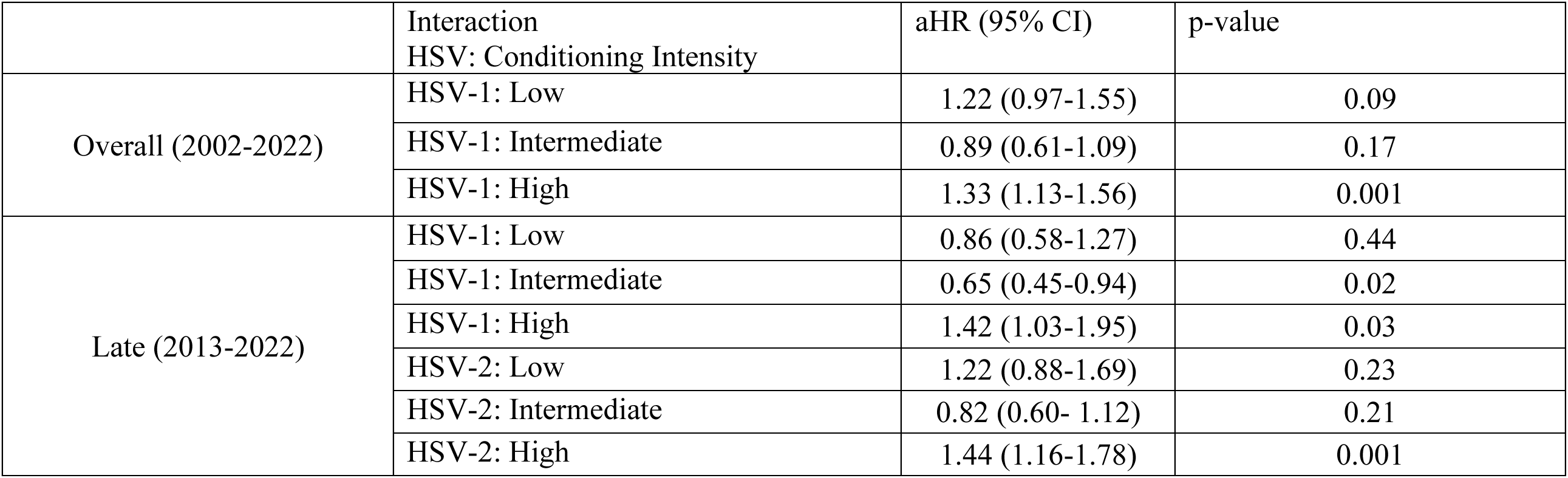
Stratum-specific HSV aHRs by conditioning intensity for significant interaction terms.

**Supplemental Figure 1:**
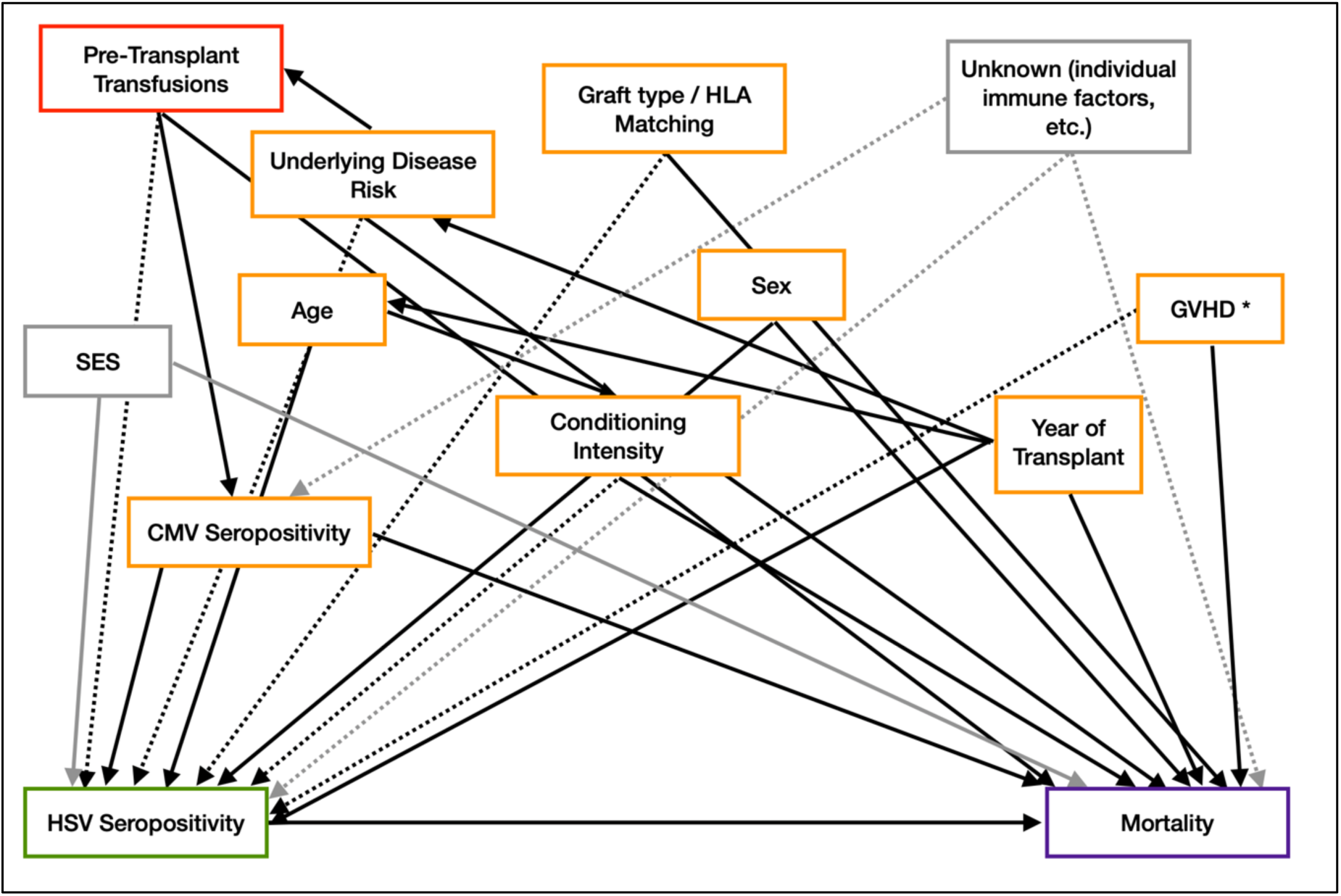
Directed acyclic graph of the relationship between HSV and other risk factors with mortality. The exposure (HSV seropositivity) is shown in green and the outcome (mortality) is shown in purple. Unmeasured or unknown factors (ex: SES) are shown in gray. Solid lines indicate known/established relationships between factors; dotted lines indicate possible associations. GVHD is starred because the directionality of the association between HSV and GVHD is unclear; i.e. GVHD may lie on the causal pathway between HSV and mortality. Pre-transplant transfusions (in red) was a potential confounder identified in a secondary analysis and was not considered in our a priori list of variables.

**Supplemental Figure 2:**
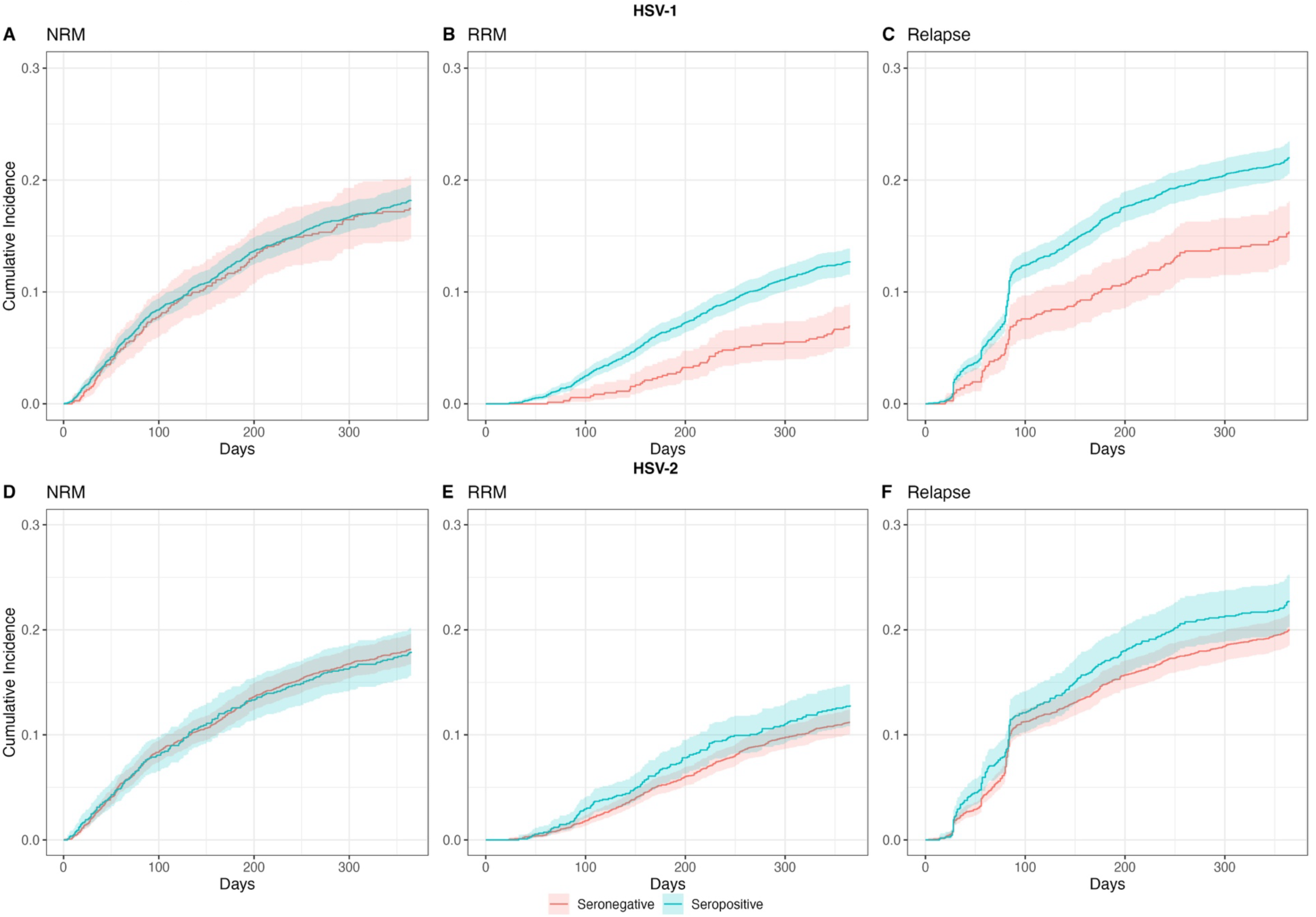
Overall (2002-2022) unadjusted cumulative incidence curves comparing NRM, RRM, and relapse by HSV-1 (A-C) and HSV-2 (D-F)

**Supplemental Figure 3:**
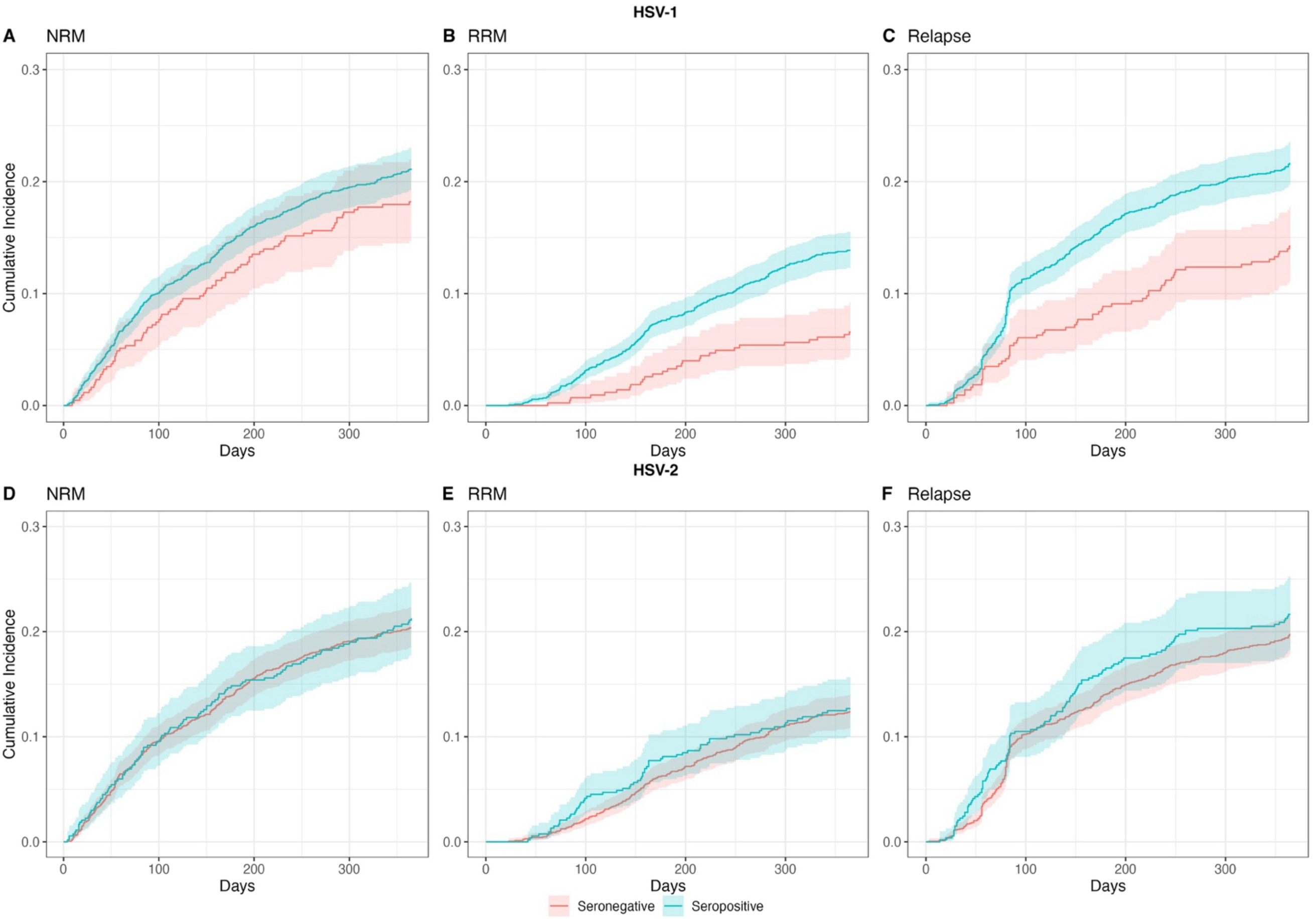
Early (2002-2012) unadjusted cumulative incidence curves comparing NRM, RRM, and relapse by HSV-1 (A-C) and HSV-2 (D-F)

**Supplemental Figure 4:**
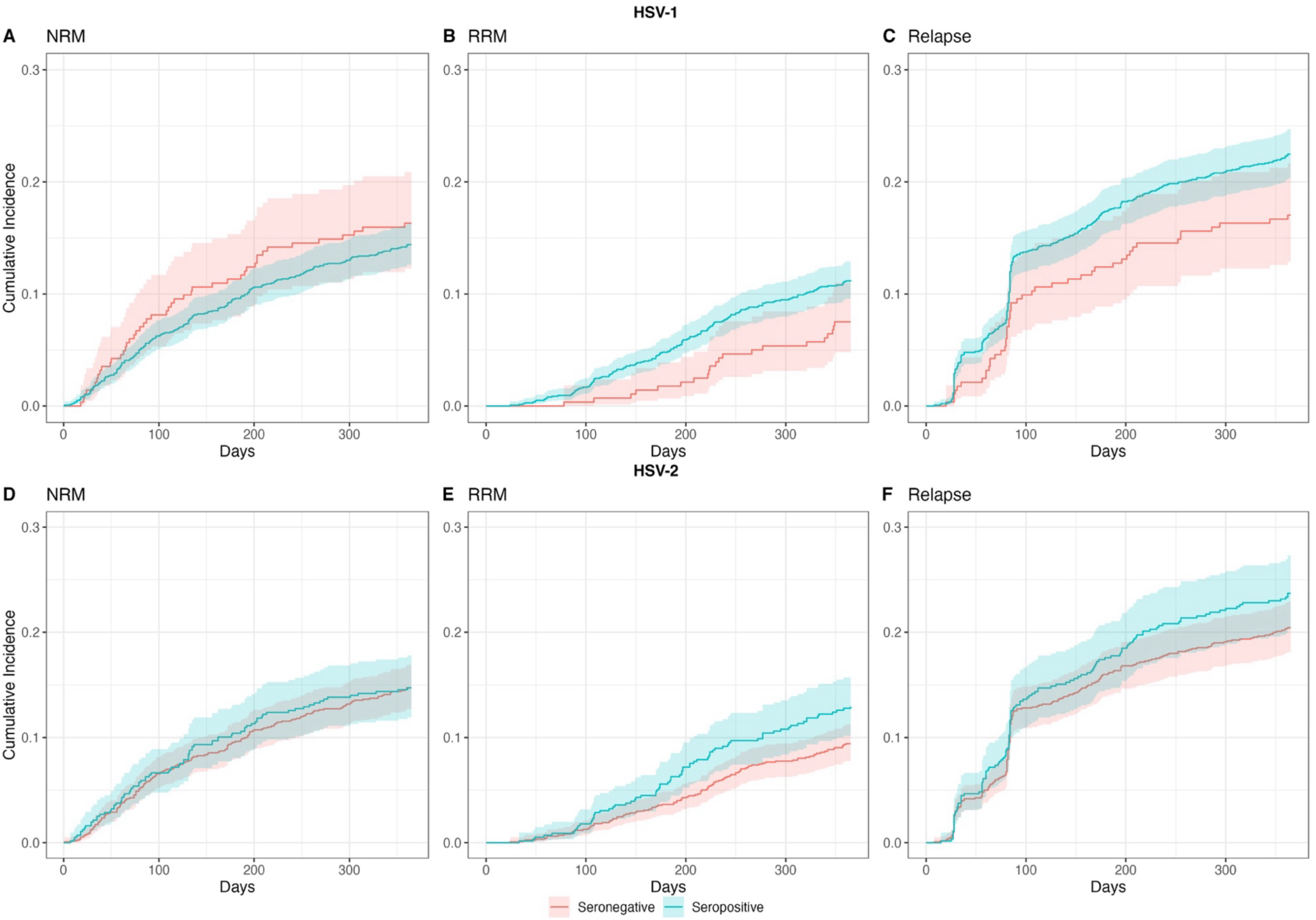
Late (2013-2022) unadjusted cumulative incidence curves comparing NRM, RRM, and relapse by HSV-1 (A-C) and HSV-2 (D-F)

